# Scalable long-read Nanopore HPV16 Amplicon-based Whole-Genome Sequencing

**DOI:** 10.1101/2025.05.22.25328149

**Authors:** Maina K. Titus, David Giesbrecht, Cliff I. Oduor, Kapten Muthoka, Karamoko Niaré, Rebecca Crudale, Abebe A. Fola, Sin Ting Hui, Isaac E. Kim, Philip K. Tonui, Peter M. Itsura, Ronald Tonui, Ann M. Moormann, Patrick J. Loehrer, Darron R. Brown, Omenge E. Orang’o, Susan Cu-Uvin, Aaron C. Ermel, Rachel A. Katzenellenbogen, Jeffrey A. Bailey

**Affiliations:** Department of Pathology and Laboratory Medicine, Warren Alpert Medical School, Brown University, Providence, RI, USA; Center for Computational Molecular Biology, Brown University, Providence, RI, US; Academic Model Providing Access to Healthcare (AMPATH) Cervical Cancer Screening Program, Eldoret, Kenya; Department of Reproductive Health and Gynecology, Moi University, Eldoret, Kenya; Indiana University School of Medicine, Indianapolis, IN, USA; Indiana University Melvin and Bren Simon Comprehensive Cancer Centre; Division of Infectious Diseases and Immunology, Department of Medicine, University of Massachusetts Chan Medical School, Worcester, MA, USA; Division of Infectious Diseases, Warren Alpert School of Medicine, Brown University, Providence, RI, USA; Department of Obstetrics and Gynecology, Aga Khan University, Medical College, Nairobi, Kenya, East Africa

## Abstract

Human papillomavirus 16 (HPV16) drives precursor cervical lesions that often progress to cervical cancer (CC). Variation within the HPV16 genome has been associated with CC risk. Here, we developed an affordable and portable amplicon-based long-read whole genome sequencing (WGS) approach using Oxford Nanopore Technologies (ONT) to investigate HPV16 genetic diversity among women in sub-Saharan African countries. Applied to a control CaSki cell line and clinical samples (n = 12), our method generated complete HPV16 genomes at high coverage (median read coverage: 5,899–15,279×). Benchmarking our HPV16 controls showed high accuracy for two variant calling pipelines (Clair3 and PEPPER-Margin DeepVariant). Phylogenetic analysis identified all four previously defined HPV16 lineages (A–D) and their high-risk sublineages. All lineages exhibited strong concordance across *de novo* assembly, reference-based phylogenetics, and unsupervised clustering. Our pipeline effectively captured the full extent of genomic variation, including putative lineage-informative SNPs. This method offers a robust amplicon-based WGS and analysis pipeline for HPV16, making it well-suited for integration into surveillance, diagnostics, and epidemiological efforts in low-resource areas.

## Introduction

Cervical cancer (CC) is a preventable and potentially curable disease; however, it remains the fourth most common cancer among women, with global estimates of 604,000 new cases and 342,000 deaths occurring annually^1^. The vast majority of these deaths occur in countries with low Human Development Index (HDI), particularly in sub-Saharan Africa^2^. Cervical cancer prevention primarily involves human papillomavirus (HPV) vaccination and screening for precursor lesions, followed by appropriate treatment. In 2020, the World Health Organization (WHO) aimed to eliminate CC by 2030, establishing the 90–70–90 targets: (1) vaccinating 90% of girls with the HPV vaccine by age 15, (2) screening 70% of women with a high-performance test at ages 35 and 45, and (3) ensuring that 90% of women with cervical pre-cancer or invasive CC have access to appropriate treatment^3^.

When implemented at scale, current screening methods effectively reduce the incidence and mortality of CC, with evidence suggesting HPV DNA detection alone is the most effective approach^4–6^. In 2021, the WHO updated its guidelines for screening and treating cervical precancerous lesions, supported by two pivotal studies that reinforced the superiority of primary HPV nucleic acid detection as a screening strategy^4,7^. Cervical intraepithelial neoplasia (CIN), a precursor to CC, is often driven by high-risk HPV (HR-HPV) infection. To date, more than 200 HPV genotypes have been identified and classified as either high-risk (HR) or low-risk (LR) based on their association with CC^8^. While WHO recognizes primary HPV testing as central to advancing CC screening^5,6^, sequencing HR-HPV genotypes offers more profound insight into disease risk, with specific lineages and sub-lineages carrying different prognostic implications^9–11^.

Human papillomavirus type 16 (HPV16) is the most common HR-HPV type globally and is responsible for over 50% of worldwide CC cases^12^. In sub-Saharan Africa, HPV16 remains the dominant HR-HPV among women, regardless of cervical lesion status^13–17^. This viral type is further divided into four major evolutionary lineages: A (formerly European type), B (African type-1), C (African type-2), and D (Asian type), with specific variant sub-lineages, such as A4 and D2, associated with an increased risk of CC^9,18–20^. While variants from Europe and Asia A lineage have been extensively characterized, HPV16 isolates from Africa remain understudied, and few have been sequenced, creating a significant knowledge gap regarding their lineage and sub-lineage, as well as their specific variations and associations with CC in sub-Saharan Africa.

Most HPV16 genomic studies in Africa have relied on targeted sequencing, focusing primarily on the oncogenic early gene E6, the major capsid late gene L1, and the non-coding long control region (LCR), leaving significant portions of the genome unexplored^21–23^. Moreover, these approaches may fail to detect integrated HPV sequences. Whole-genome sequencing (WGS) offers a comprehensive landscape of HPV16 genetic diversity, which has been challenging to achieve with short-read sequencing (e.g., Illumina) platforms and relatively expensive to implement in low-resource settings like sub-Saharan Africa. The recent development and maturation of long-read Oxford Nanopore Technologies (ONT) sequencing presents a promising alternative, offering portable, cost-effective, and real-time sequencing solutions through platforms such as the MinION and PromethION2 (P2).

These technologies have been successfully deployed for pathogen genomic surveillance, including Tuberculosis, SARS-CoV-2, and Ebola virus, demonstrating the potential for improving genome assembly and variant detection through this long-read sequencing (LRS)^24–26^. Given its affordability and adaptability, ONT sequencing holds promise for fully covering the genome in one or a few amplicons, thereby expanding HPV16 WGS in sub-Saharan Africa, where the burden of CC is highest.

Despite the increasing adoption of Nanopore LRS, no universally accepted variant caller exists, and it continues to be an area of active development. This differs from short-read sequencing, where the Genome Analysis Toolkit (GATK) and other variant callers are well-established as sensitive and specific^27,28^. Several tools are available for ONT variant calling, including Medaka, Clairvoyante, Clair, and Nanocaller, which utilize pileup-based methods ^29,30^, while PEPPER employs a full- alignment approach^29^. It maximizes precision and recall through full alignment. Clair3, the successor to Clair, merges pileup-based and full-alignment strategies to enhance efficiency and accuracy^30^.

These variant callers are typically benchmarked using well-characterized human genome datasets from the Genome in a Bottle (GIAB) consortium rather than viruses or other organisms^31^. Thus, it is essential to determine optimal parameters and validate which program has the best recall and precision for HPV. It requires proper controls that are already accurately sequenced and assembled (aka a “truth set”). We aimed to establish an HPV16 ’truth set’ variants dataset to benchmark PEPPER and Clair3, two of the most widely used long-read variant callers.

We developed a scalable HPV16 WGS and analysis pipeline using ONT’s MinION and P2 platforms. By combining multiple primer sets for long-range PCR, including both tiling and full-length, we aimed to traverse the entire genome and potentially capture the circular form. We applied this approach to the HPV16-positive CaSki cell line, clinical isolates from western Kenya, and clinical isolates from Rwanda. To benchmark variant calling accuracy, we established a high-confidence HPV16 “truth set” of variants and evaluated PEPPER and Clair3, both of which exhibited strong performance for whole-genome variant detection. Our method achieved complete genome recovery with high coverage, enabling accurate phylogenetic classification into lineages A, B, C, and D. This robust and adaptable HPV16 WGS workflow facilitates comprehensive genomic surveillance and diagnostics, particularly in resource-limited settings.

## Results

### WGS primer design to capture HPV16

To comprehensively analyze the genetic diversity of HPV16, we developed a long-range PCR viral enrichment strategy that leveraged three primer set designs for amplification. The primer designs were (1) a near full-length primer set generating amplicons up to 7.7 kb to capture intact or nearly full- length HPV16 DNA (complete genome measures ∼ 8 kb), (2) a tiling-set of three primer pairs yielding overlapping amplicons of 2.1 kb, 3.9 kb, and 2.6 kb, (**Fig. 1a**) and a junction primer pair designed to capture the genome undergoing linearization and integration into the host DNA, as illustrated in (**Fig. 1b)**. This last primer pair addresses HPV16’s episomal form and tandem integration into the host genome (extrachromosomal form)^32^ (**Fig. 1b**). We first tested our primers using DNA extracted from the HPV 16-positive, cervical cancer CaSki cell line and sequencing amplicons with the Oxford Nanopore MinION sequencer. Our method was sensitive, allowing us to amplify and sequence as few as five copies of HPV16 per reaction in 2 μL of CaSki DNA input (**Supplementary** Fig. 1a **and Supplementary** Fig. 1b). Our analysis workflow (**Fig. 1c**) enabled us to accurately align the amplicon sequences to the expected region of the HPV16 reference genome (GenBank: K02718.1) (**Supplementary** Fig. 1c). We pooled and sequenced all PCR products to ensure sufficient coverage for accurate variant calling and other downstream analyses. We achieved a deep (>1000X) read depth across the HPV16 genome from the CaSki cell line DNA **(Fig. 1d** **and Supplementary** Fig. 1d).

**Figure 1.**
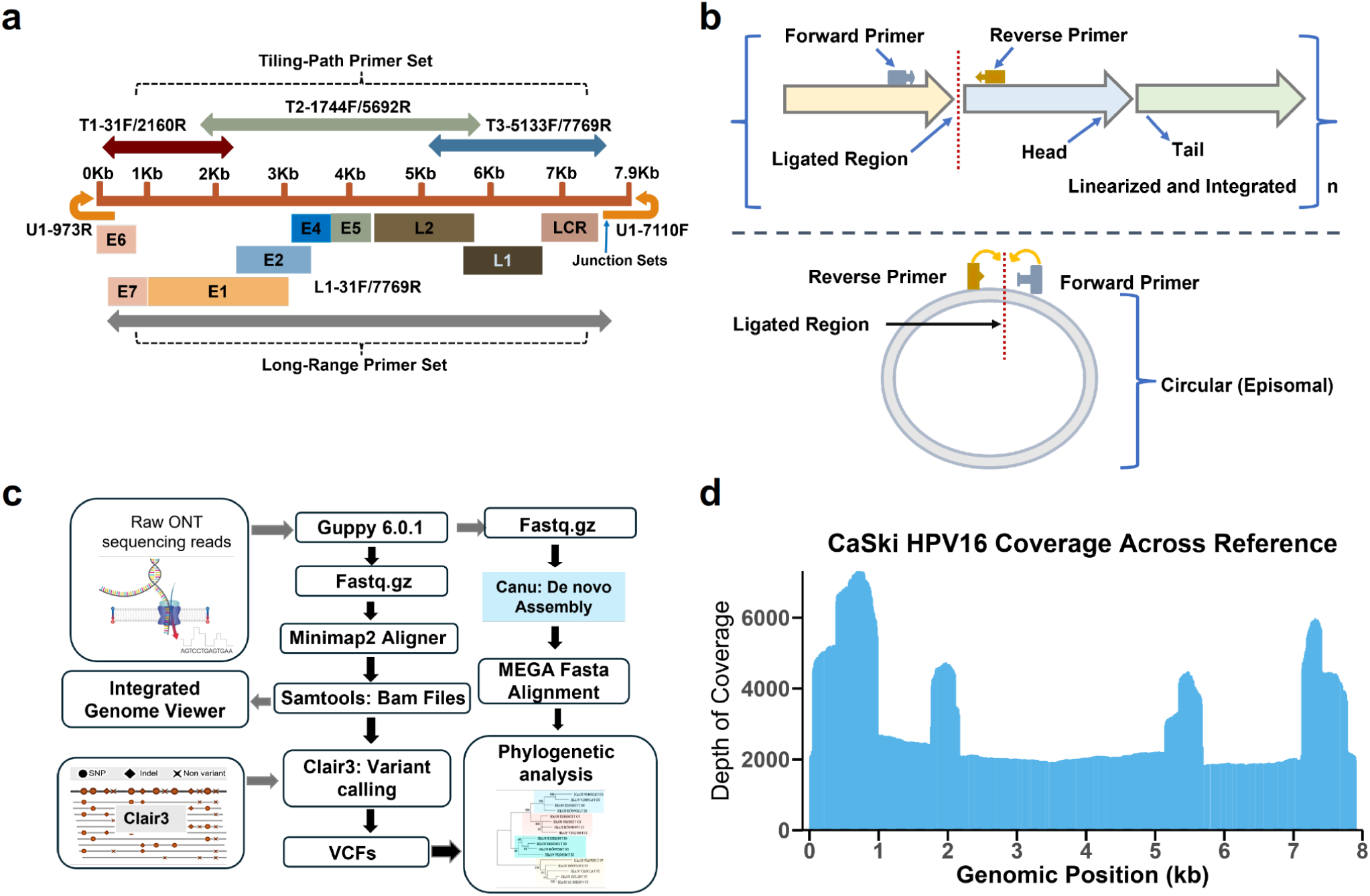
| A trio primer set for HPV16 whole-genome sequencing enrichment. **(a)** Schematic of the primer design for viral pre-sequencing enrichment. On top of the panel, the tiling-path primers are set: T1 (dark red), T2 (light green), and T3 (blue), along with the U1 (yellow) primer set, which is essential for amplifying the ligated region at the genome’s terminus. At the bottom of the panel is the near full-length primer set (gray). **(b)** Representation of HPV16 in its integrated extrachromosomal form as a multi-tandem repeat (upper) and episomal form (lower). **(c)** HPV16 long-read analysis pipeline. ONT reads were base-called with Guppy, aligned using Minimap2, and processed with SAMtools. Variants were called with Clair3. De Novo Assembly was executed using Canu, followed by sequence alignment and phylogenetic analysis in MEGA. **(d)** Representative depth-of-coverage plot showing sequencing read depth across the CaSki HPV16 genome relative to the reference for combined amplicons.

### Benchmarking PEPPER and Clair3 for HPV16 WGS Variant Calling

To assess the accuracy of long-read variant calling, we benchmarked HPV16 controls comparing PEPPER and Clair3 variant calling against GATK4 (Methods). We evaluated the performance of the variant callers using precision, recall (also known as sensitivity), and F1-score (the harmonic mean of precision and recall). Among the 29 consensus variants identified by all three callers, Clair3 and GATK4 jointly detected two additional variants that PEPPER missed **(Fig. 2a)**. Meanwhile, PEPPER and GATK4 identified one variant that Clair3 missed. Additionally, PEPPER identified one variant that was not detected by either Clair3 or GATK4 **(Fig. 2a)**. Overall, Clair3 achieved 100% precision, 96.9% recall, and a 98.4% F1-score, whereas PEPPER attained 96.8% precision, 93.8% recall, and a 95.2% F1-score **(Fig. 2b**; **Table 1)**.

**Figure 2.**
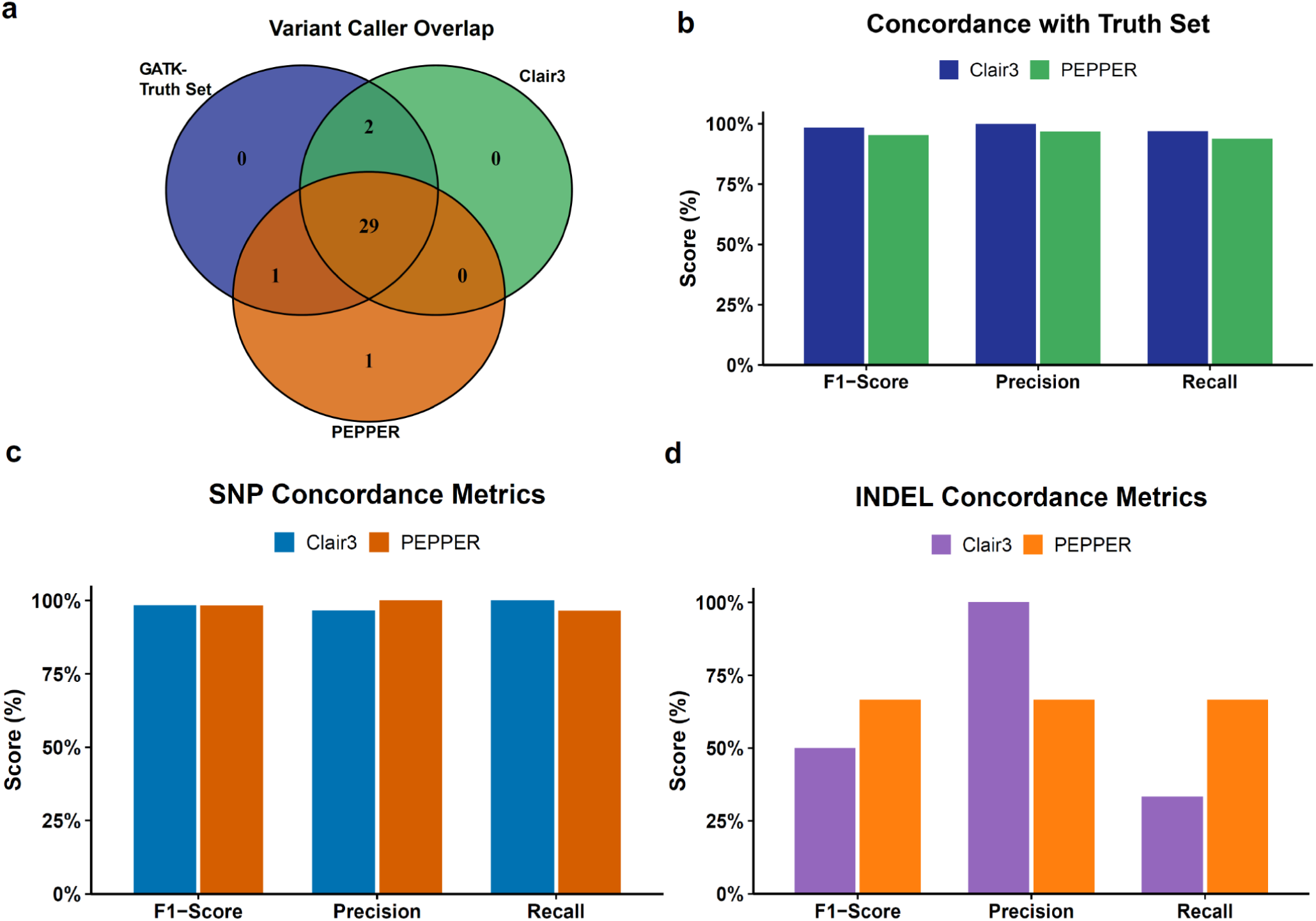
| Performance comparison of Clair3 and PEPPER against GATK. **(a)** A Venn diagram shows the overlap among the variant callers: PEPPER (brown), Clair3 (green), and the Truth-set GATK. **(b)** Overall concordance performance of Clair3 (blue) and PEPPER (green) relative to the Truth-set GATK, with F1-Score, Precision, and Recall on the x-axis and percentages on the y-axis. **(c)** SNP concordance metrics for Clair3 (blue) and PEPPER (brown), displaying F1-Score, Precision, and Recall on the x-axis and percentages on the y-axis. **(d)** INDEL concordance metrics for Clair3 (purple) and PEPPER (yellow), featuring F1-Score, Precision, and Recall on the x-axis and percentages on the y-axis.

**Table 1.**
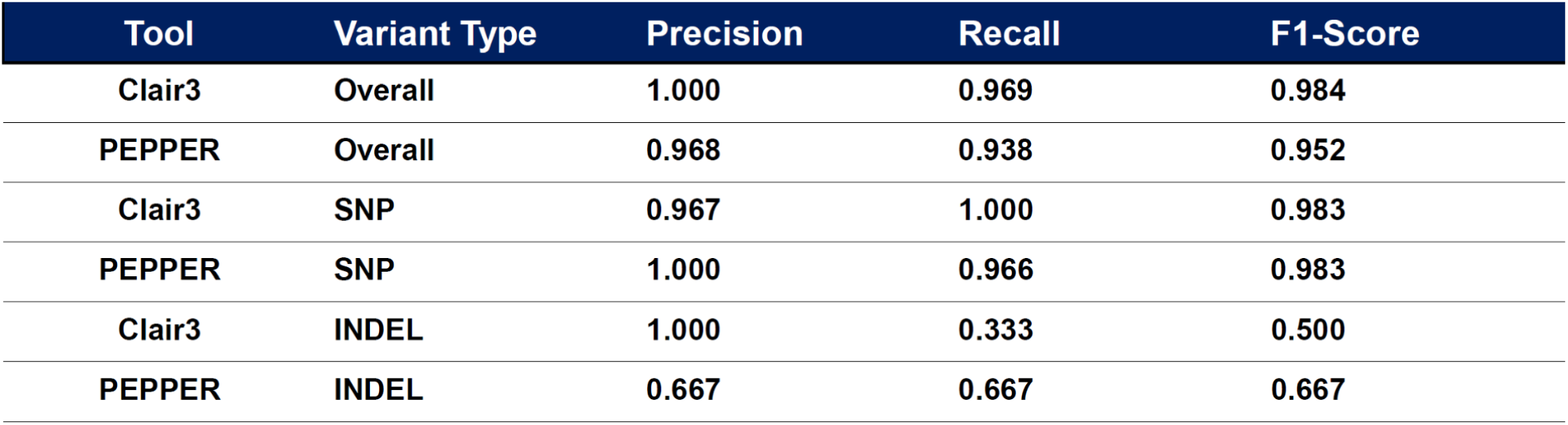
| Performance comparison of Clair3 and PEPPER Margin DeepVariant (PEPPER) with GATK4. The table summarizes the performance metrics of Clair3 and PEPPER in relation to the GATK truth set. It presents overall performance alongside specific metrics for SNPs and INDELs, providing precision, recall, and F1-score values for each variant type.

When assessing SNPs, Clair3 achieved 96.7% precision and 100% recall, while PEPPER attained 100% precision and 96.6% recall, resulting in equivalent F1 scores of 98.3% **(Fig. 2c**; **Table 2)**. For indel detection, Clair3 exhibited 100% precision but only 33.3% recall, leading to an F1 score of 50% due to its conservative calling. In contrast, PEPPER achieved 66.7% precision, 66.7% recall, and a 66.7% F1 score **(Fig. 2d**; **Table 2)**. These findings are consistent with known challenges of Oxford Nanopore Technologies (ONT) sequencing, particularly with R9.4.1 flow cells and Guppy basecalling models, where small indel errors frequently occur, particularly in homopolymer regions^33^.

**Table 2.**
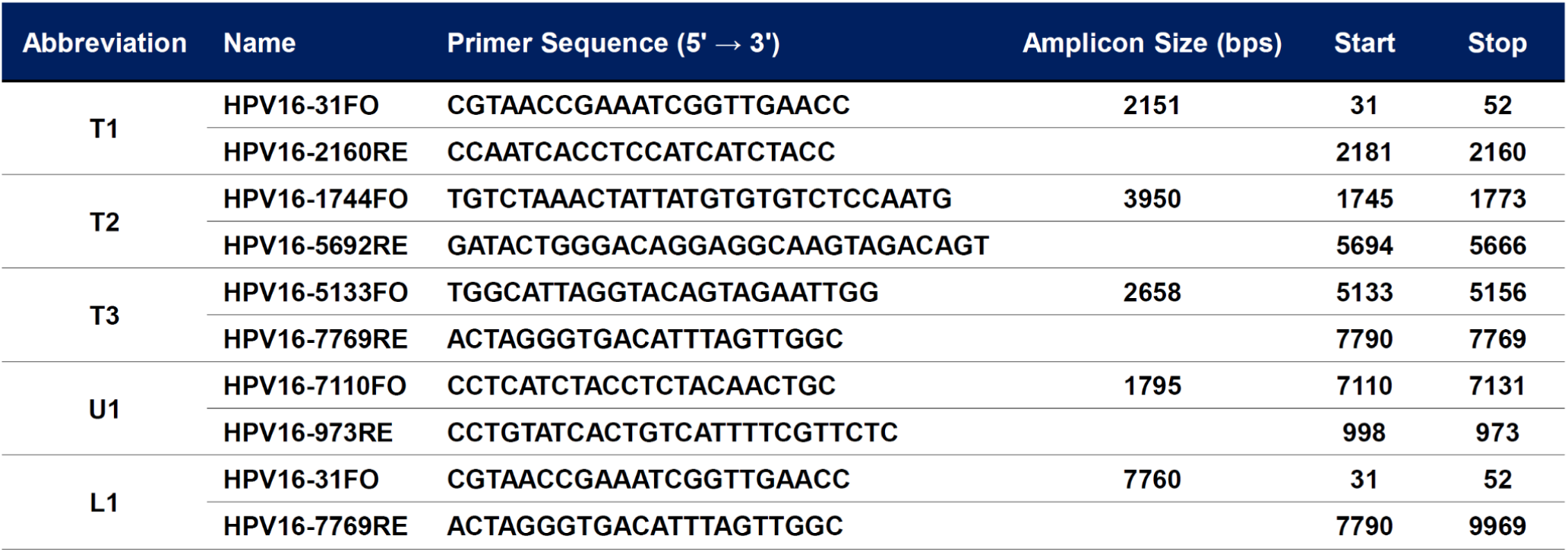
| Primer pairs used in this study for complete whole-genome sequencing of HPV16. Primer names, abbreviations, and corresponding sequences are presented alongside their nucleotide start and stop positions. All coordinates provided relative to the HPV16 reference genome (GenBank accession: K02718.1).

### HPV16 WGS of clinical samples

Next, we evaluated the performance of our approach on 12 HPV16-positive isolate samples extracted from cervical exfoliated cells and genotyped using the Roche Linear Array (LA). These clinical samples were obtained in previous studies from women in western Kenya (n=10) and Rwanda (n=2) who participated in cervical cancer-related studies and were archived for future research^14,34^. Libraries for the samples were prepared, barcoded, and sequenced using the ONT Rapid Barcoding kit (SQK- RBK110.96, Oxford Nanopore). All samples yielded a good depth of coverage (median read coverage ranging from 5,899 to 15,279) **(Fig. 3; Supplementary** Fig. 2), and 100% complete genome coverage was observed in clinical samples. The depth exceeded 500X in regions that had previously posed challenges with Illumina sequencing^20^, including E2, E4, E5, and L2 **(Fig. 3)**. Given the use of overlapping primer pairs, we observed peaks in the sequencing data, reflecting the overlapping regions of the amplicons across all samples **(Fig. 3)**. This pattern consistently appeared in the HPV16 coverage bar plots, regardless of lineages **(Fig. 3),** demonstrating robust and uniform coverage across samples.

**Figure 3.**
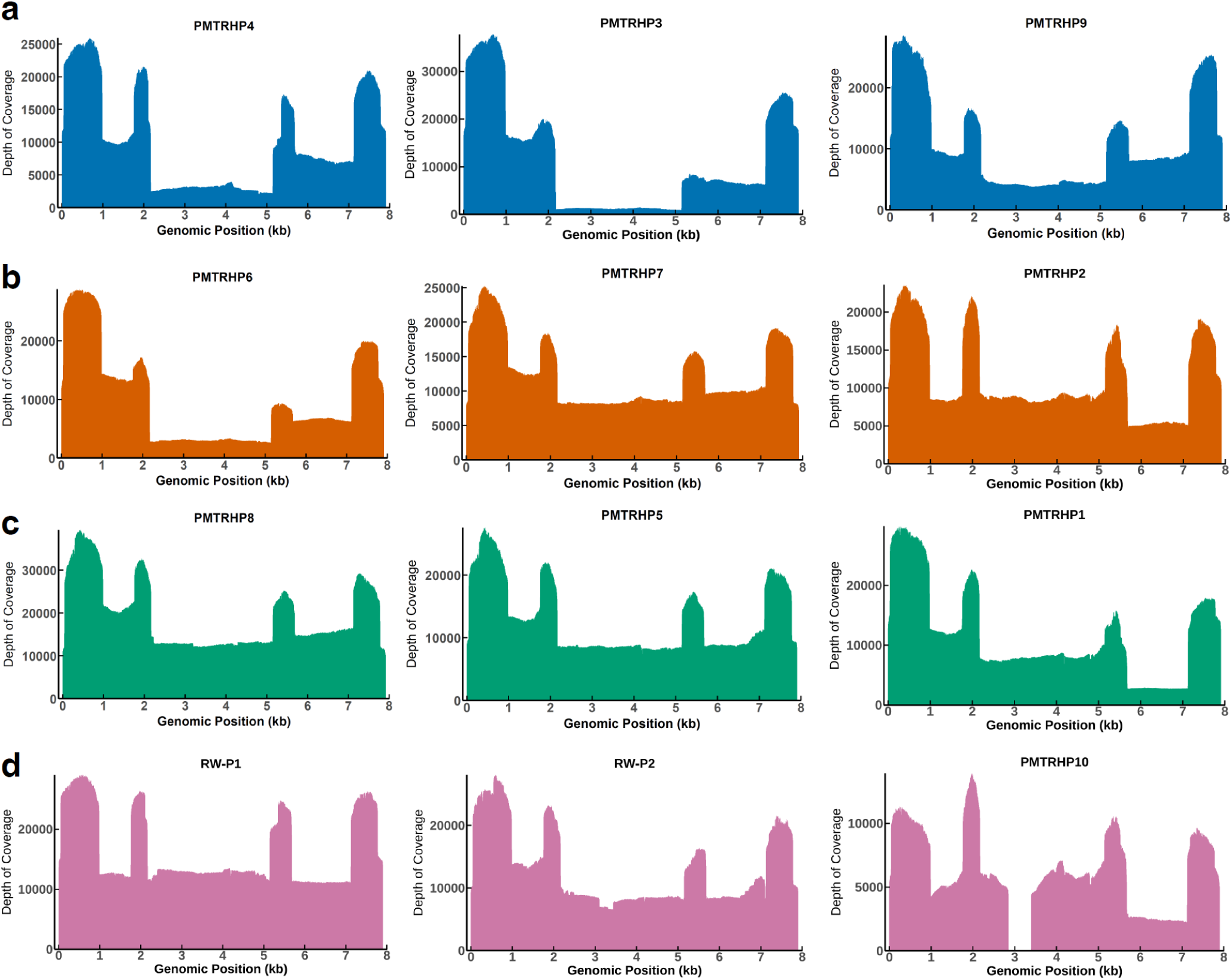
| Depth of coverage analysis of HPV16 genomes in clinical cervical isolates. Read depth and coverage profiles for 12 HPV16-positive clinical samples, categorized by lineage, were generated from bedgraph files. **(a)** Lineage A (blue), **(b)** Lineage B (vermilion), **(c)** Lineage C (green), and **(d)** Lineage D (magenta), with each panel representing three clinical samples per lineage. The x-axis indicates the genomic position (kb) of HPV16, and the y-axis shows the sequencing depth. Sample PMTRHP3 exhibited greater than 1,000× coverage, while PMTRHP10 had greater than 500× coverage in a reduced depth region. High-intensity peaks correspond to overlapping amplicon regions, demonstrating strong and consistent coverage across samples.

### Robust recovery and phylogenetic analysis of HPV16 genomes from clinical samples

Next, we evaluated the compatibility of sequencing data from clinical samples (n=12) generated by our WGS approach for downstream analyses, including *de novo* assembly and reference-based variant calling. Reference genomes for HPV16 variant lineages and sublineages were retrieved from GenBank and merged with *de novo*–assembled sequences generated using the canu package. Molecular evolutionary genetic analysis software (MEGA) was employed for pairwise and multiple sequence alignments, followed by maximum likelihood phylogenetic tree construction.

Our method successfully recovered complete HPV16 genomes from all clinical samples **(Fig. 4a)**. Phylogenetic analysis revealed four distinct lineages: A, B, C, and D **(Fig. 4a)**. Within lineage A, two samples were classified as sublineage A1 and one as A2. Samples from lineage B aligned with sublineage B4, Lineage C with sublineage C1, and Lineage D samples with sublineage D3. These results demonstrate that our WGS method reliably recovers high-quality complete HPV16 genomes, enabling precise phylogenetic classification.

**Figure 4.**
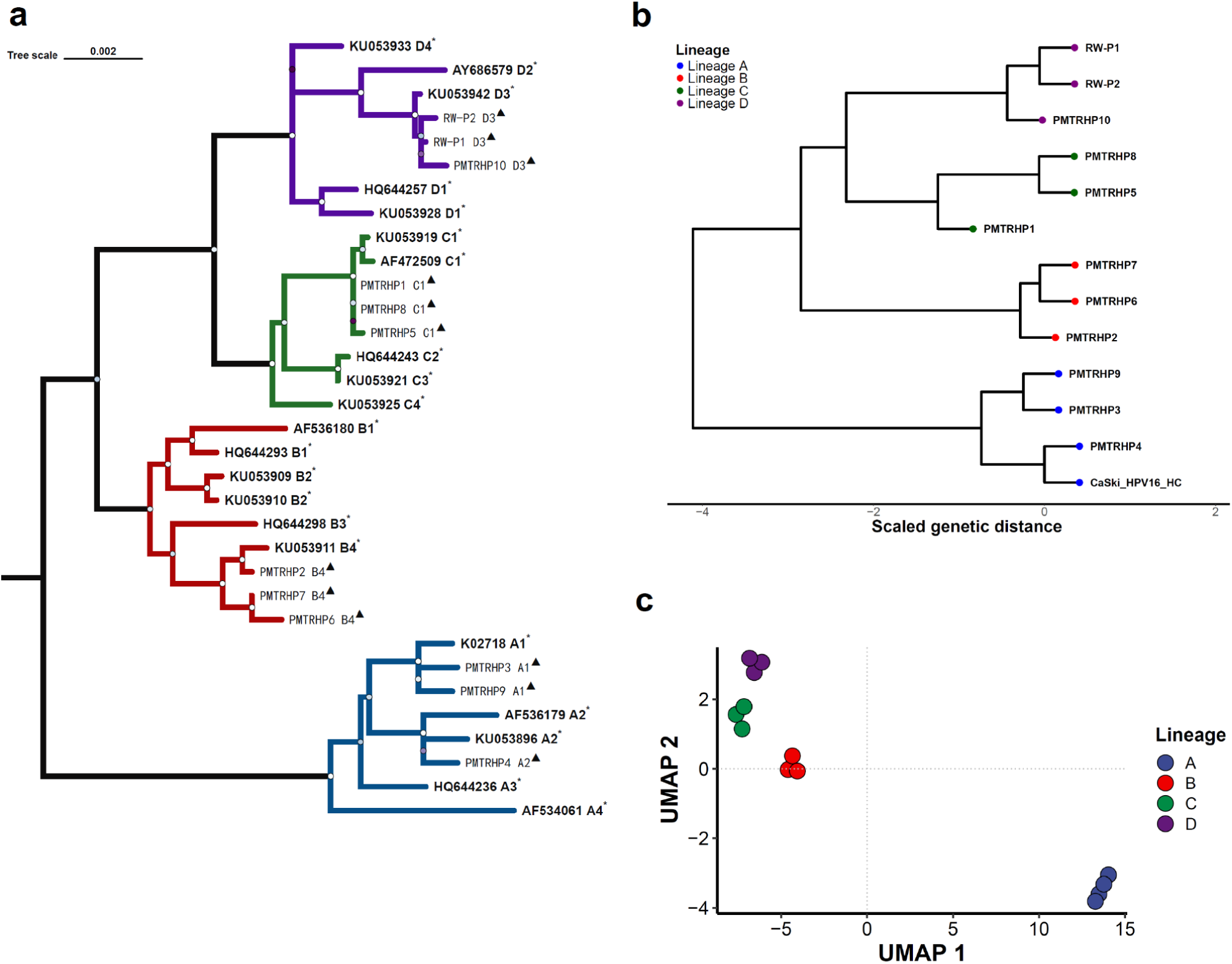
| Phylogenetic tree and UMAP clustering of HPV16 genomes. **(a)** A maximum likelihood phylogenetic tree constructed from reference HPV16 genomes (GenBank; marked with an asterisk) and assembled clinical isolates from cervical swabs (black triangles). Branch colors indicate the four major HPV16 lineages. The scale bar represents genetic distance. Node support is visualized with gradient circles, where brighter shades indicate higher bootstrap values. **(b)** A phylogenetic tree generated from variant call format (VCF) files, with tip colors corresponding to HPV16 lineages as in panel a. **(c)** A UMAP plot based on processed VCF data, illustrating unsupervised clustering of HPV16 genomes. In all panels, HPV16 lineages are color-coded as follows: A (blue), B (red), C (green), and D (purple).

In parallel, reference-based alignment and variant calling were performed on sorted BAM files using Clair3, with variant call format (VCF) files subsequently merged via BCFtools. A custom bash script and R workflow were used to process VCFs for phylogenetic tree construction and Uniform Manifold Approximation and Projection (UMAP) clustering **(Fig. 4b, 4c)**. As expected, the reference-based phylogeny reproduced the four lineage clusters (A, B, C, and D) **(Fig. 4b)** observed in the *de novo* analysis **(Fig. 4a)**. UMAP clustering further confirmed these four distinct groups **(Fig. 4c)**.

The concordance across *de novo* assembly, reference-based phylogeny, and unsupervised clustering underscores the robustness of our method for recovering complete HPV16 genomes. This method enabled precise lineage classification and downstream evolutionary analyses, affirming the extensive genetic heterogeneity of HPV16 as observed in previous studies^10,35,36^.

### Mutational profiles of clinical HPV16 isolates

To further evaluate the suitability of ONT WGS and our optimized bioinformatics pipeline for HPV16 genomic analysis, we analyzed viral genomes from our clinical samples using the PaVe HPV16 reference (accession number K02718). Across all samples, we identified a total of 253 variants, the vast majority of which were SNPs (244/253, 96.4%), while the remainder was insertions and deletions (indels; 9/253, 3.6%), including two insertions and seven deletions **(Fig. 5a)**. This corresponded to an average of one variant per 31 nucleotides. When classified by predicted functional impact, 52.8% of the variants were synonymous and non-coding SNPs, and 47.2% were missense mutations, resulting in a missense-to-silent ratio of 0.89 **(Fig. 5b)**. The lack of stop codons supports the accuracy of the sequencing since these clinical isolates should be intact. Excluding the highly variable upstream regulatory region (URR), variants were broadly distributed across the remaining coding regions **(Fig. 5c)**, with the following gene-level counts: E1 (n = 44), E2 (n = 37), E5α (n = 12), E6 (n = 14), E6 (n = 7), E7 (n = 4), E8^E2 (n = 26), L1 (n = 28), and L2 (n = 56). Regarding normalized variant frequency by gene, E5 exhibited the highest number of variants, followed by L2 and E2. At the same time, E7 was the least variable gene^37,38^ **(Fig. 5c)**. Focusing specifically on missense mutations, E2 harbored the highest number (n = 24), followed by E1 and L2 (n = 17). In contrast, E7 contained only a single missense SNP **(Fig. 5d)**.

**Figure 5.**
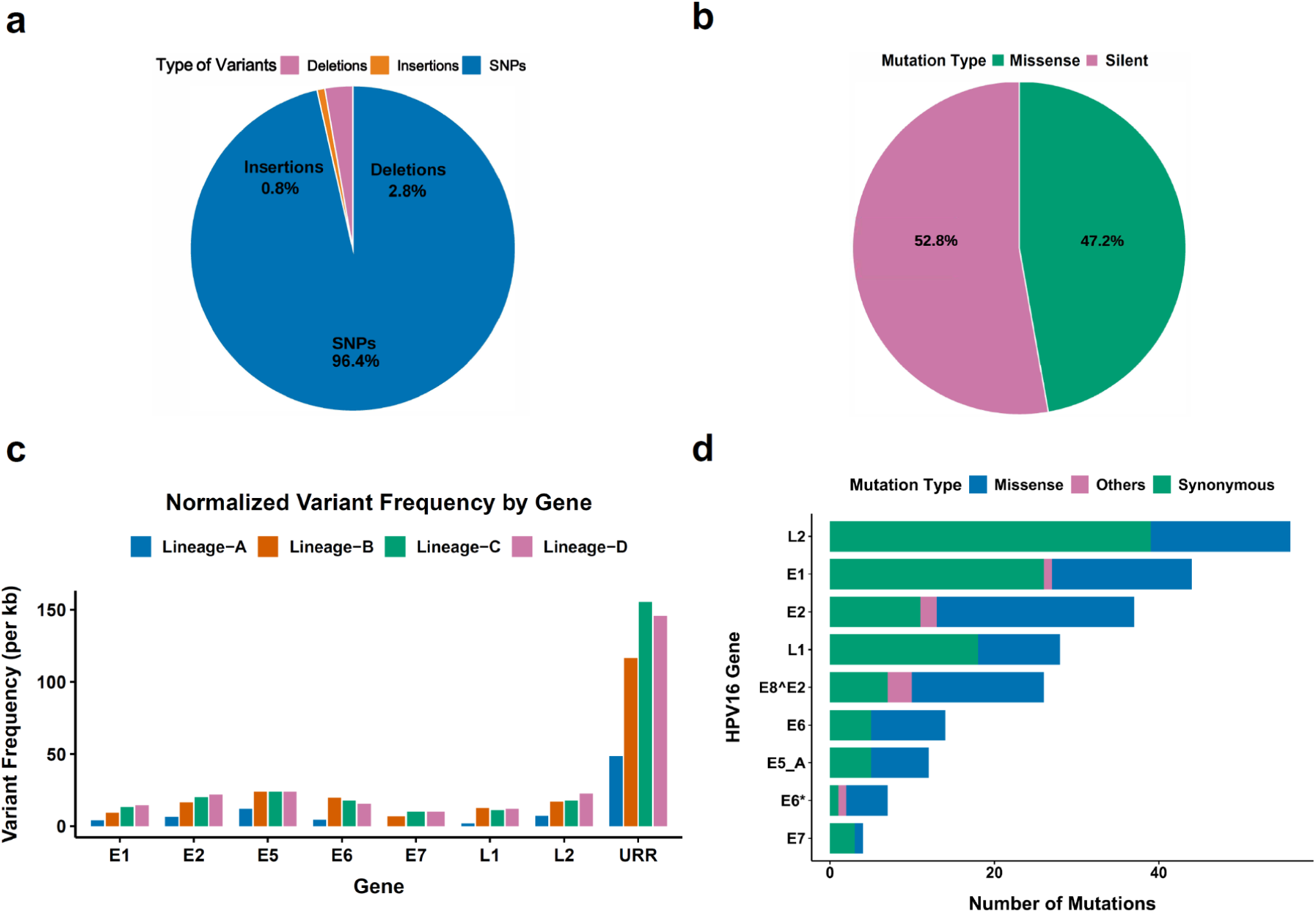
| Analysis of fixed mutations in HPV16 clinical isolates. **(a)** Distribution of variant types: single-nucleotide polymorphisms (SNPs; blue), insertions (vermilion), and deletions (magenta). **(b)** Ratio of missense (green) to silent (magenta) mutations. **(c)** Normalized variant frequency by gene across HPV16 lineages, color-coded as A (blue), B (vermilion), C (green), and D (magenta). **(d)** Mutation counts across HPV16 genes: synonymous (green), missense (blue), and other protein- altering mutations (magenta) such as disruptive in-frame, frameshift, and stop-gained variants.

## Discussion

Here, we present a robust amplicon-based WGS approach for HPV16, employing a trio-primer enrichment strategy optimized for ONT. This method achieves full genome coverage, including previously challenging regions such as E5, E2, and URR^20^, and performs well with low-abundance samples. Junction-specific primers enable complete genome recovery in circular or tandem forms. The method leverages ONT platforms, including PromethION 2 Solo (P2) and MinION, which are well- suited for deployment in resource-limited settings. A previous study employed the MinION for HPV- ICB2 WGS^39^ but required rolling circle amplification (RCA) prior to long-range PCR, a limitation for integrated HPV types such as HPV16 in CaSki cells, where circular templates are absent. The ONT Rapid Barcoding Kit enables cost-effective multiplexing of up to 96 samples. Our workflow begins with the collection of cervical swabs, followed by DNA extraction, PCR-based viral enrichment, and library preparation using this kit. Sequencing is performed on a single flow cell, with the total cost, from extraction to sequencing, estimated at approximately USD $25 per sample. Costs may be further reduced by repurposing the flow cell for additional sequencing runs targeting taxonomically unrelated organisms, thereby minimizing the risk of barcode cross-reactivity. Additional scalability could be achieved through the incorporation of unique molecular identifiers (UMIs), as demonstrated by Karst et al.,^40^ which may enhance high-throughput sequencing, an area historically dominated by Illumina platforms that offer high base-call accuracy (∼0.1% error) but are limited to short (∼500 bp) amplicons^41^. This scalable and cost-effective strategy has strong potential for advancing HPV16 genomic surveillance.

Applied to cervical clinical isolates previously genotyped using the Roche Linear Array, our method demonstrated complete concordance, highlighting its reliability for HPV16 detection. Comprehensive genome coverage was attained across all samples, with median read depths ranging from 5,899× to 15,279×, and minimum depths exceeding 500× even in challenging genomic regions such as E2, E4, E5, and L2. The high and uniform coverage enabled accurate de novo assembly and high-confidence variant calling, resulting in the recovery of complete, high-quality HPV16 genomes from clinical samples. Both reference-based and de novo analyses consistently identified four major phylogenetic clusters corresponding to the established HPV16 lineages: A (European), B (African 1), C (African 2), and D (Asian)^9,10,35,36^. These lineage assignments were further confirmed by unsupervised clustering using UMAP, which preserves local and global data structures, revealing a clear separation of the four groups. The precise classification of samples into their respective sub-lineages aligns with previous phylogenetic studies^9,10,35,36^, validating the accuracy and strength of our approach. The concordance across de novo assembly, reference-guided phylogeny, and dimensionality reduction-based clustering underscores the robustness and resolution of our method and analysis pipeline.

Importantly, our approach significantly enhances accurate lineage assignment, enabling downstream evolutionary and epidemiological analyses that capture the extensive genetic heterogeneity of HPV16. The ability to resolve lineage diversity with high confidence is critical for molecular epidemiology, public health surveillance, and studies on lineage-specific pathogenicity. Furthermore, our pipeline’s capability to generate phylogenies and UMAP or t-SNE clusters directly from processed VCF files offers an alternative and scalable framework for HPV16 genotyping, eliminating the need for full- genome assemblies. Additionally, our method effectively captured the full extent of genomic variation, with one variant occurring approximately every 31 nucleotides, predominantly as SNPs.

Coding changes were evenly split between synonymous and missense mutations (ratio = 0.89). Variants were broadly distributed, showing enrichment in E5, L2, and E2, while E7 remained highly conserved, consistent with prior reports^37,38^. Collectively, these findings establish our WGS workflow as a reliable and high-resolution platform for HPV16 genomic analysis, with broad utility in clinical diagnostics, viral evolution studies, and public health surveillance.

PEPPER and Clair3 variant calling demonstrated strong overall performance in HPV16 whole-genome variant analysis. Both tools achieved comparable F1-scores and exhibited high precision and recall for SNP detection, highlighting their suitability for single-nucleotide variant analysis in viral genomes.

However, indel calling remained a challenge, with reduced performance across both callers. This limitation reflects well-documented constraints of ONT data, particularly from R9.4.1 flow cells and Guppy basecalling, where systematic errors are prevalent in homopolymeric and repetitive regions^31,33^. These platform-specific biases underscore the need for improved indel resolution and benchmarking strategies tailored to ONT-based viral genome analysis. Our study integrated ONT data with publicly available Illumina data from CaSki cell lines; however, minor differences in cell line sources may have slightly affected indel concordance. Future comparisons should utilize matched samples to enhance evaluation consistency. As ONT technology continues to evolve, gains in basecalling accuracy and chemistry are expected to enhance variant detection. Both callers will likely benefit, improving their utility for high-resolution HPV16 genomic studies. Importantly, the tools employ distinct computational strategies. PEPPER uses full alignment, while Clair3 combines pileup-based and full alignment approaches, offering different trade-offs between accuracy and computational efficiency. These considerations are critical when scaling analyses across large cohorts. Overall, this work establishes a framework for future investigations into HPV16 genomic diversity and underscores the need for validation in diverse, independent sample sets to support broader clinical and epidemiological applications.

This study has limitations. We did not quantify HPV16 viral load by qPCR to correlate with sequencing depth; instead, we relied on a Qubit estimate for known CaSki cell copy numbers, which approximate a detection limit of five copies per 2 μL. While the method employs three singleplex reactions, minimal benefit is anticipated from two-tube multiplexing. As a proof of concept, future work should focus on primer optimization to enable single-tube multiplexing, thereby improving scalability and reducing costs. INDEL detection remained suboptimal due to known ONT limitations with R9.4.1 flow cells and Guppy basecalling in homopolymeric regions. However, advances in Kit 14 chemistry, R10.4.1 flow cells, and improved models are expected to enhance variant calling accuracy.

In conclusion, we have developed a scalable and deployable WGS workflow for both linear integrated and episomal HPV16 using ONT long-read technology, coupled with an optimized analysis pipeline. We generated a benchmark set of HPV16 ’truth set’ variants and demonstrated that PEPPER and Clair3 variant callers perform reliably for SNP detection, supporting their potential utility in HPV16 variant analysis. While further validation in larger, diverse cohorts is warranted for broader clinical adoption, the strong concordance observed across de novo assembly, reference-based phylogenetics, and unsupervised clustering highlights the robustness of our approach for reconstructing complete HPV16 genomes. A forthcoming study will leverage this strategy to investigate the epidemiological implications of HPV16 in sub-Saharan Africa. Notably, the capacity of our WGS framework to resolve fine-scale variation across the HPV16 genome enables robust analyses of coding diversity and its potential functional implications. This work contributes to global efforts in HPV surveillance and CC elimination, particularly in high-burden, resource-limited settings.

## METHODS

### Study design, population, and ethical approvals

Ten archived cervical swab specimens were obtained from women enrolled at cervical cancer screening centers under the Academic Model Providing Access to Healthcare (AMPATH) in western Kenya, and two from the Einstein-Rwanda Research and Capacity Building Program (ER-RCBP) at Research for Development (RD Rwanda), Rwanda. These samples were collected as part of ongoing studies approved by the US National Cancer Institute (NCI) and the National Institutes of Health (NIH). Written informed consent was obtained from all participants. Ethical approvals were obtained from the Institutional Research and Ethics Committee (IREC) of Moi Teaching and Referral Hospital (MTRH) and Moi University, Eldoret, Kenya (reference number IREC/371/2022), as well as from the Institutional Review Board (IRB) of Brown University, Providence, Rhode Island, USA (IRB-ID: 2023003553). Additionally, this study utilized the HPV16-positive CaSki and HPV16-negative C33A cervical carcinoma cell lines, which were provided by the laboratory of Dr. Rachel A. Katzenellenbogen at Indiana University School of Medicine in Indianapolis, IN, USA.

## Human papillomavirus 16 detection and genotyping

### Roche Linear Array

DNA extraction from archived cervical swab specimens was performed using the Qiagen Mini-Elute kit, following the manufacturer’s instructions. Subsequently, typing was conducted using the Roche Linear Array® (LA) (Roche, Branchburg, NJ, USA) to detect 37 HPV types, including 12 high-risk, eight probable high-risk, and 17 low-risk HPV genotypes. Genotyping with LA was performed as previously described^42^, amplifying 450-bp fragments from the L1 region of the HPV virus by PCR and using a reverse line blot system for simultaneous detection of up to 37 HPV genotypes (i.e., 6, 11, 16, 18, 26, 31, 33, 35, 39, 40, 42, 45, 51, 52, 53, 54, 55, 56, 58, 59, 61, 62, 64, 66, 67, 68, 69, 70, 71, 72, 73, 81, 82, 83, 84, IS39, and CP6108). The LA assay, widely used in epidemiological studies, is regarded as the reference standard for HPV genotyping.

### DNA purification for long-range PCR and downstream next-generation sequencing

The HPV16-positive cervical swab isolates were selected for Nanopore HPV16 whole genome sequencing. The DNA for the downstream HPV16 WGS was purified from the CaSki cell line and cervical exfoliate cells. Cervical exfoliated cells were first eluted in 1 ml of 1× phosphate-buffered saline (PBS). Subsequently, DNA extraction was performed using 450 µL of the elute with the Monarch® Genomic DNA Purification Kit (New England Biolabs, Ipswich, MA, USA), following the manufacturer’s instructions. This DNA purification kit yields an excellent amount of input material for applications such as long-range PCR and downstream next-generation sequencing (NGS) library preparation. The DNA extracted from the HPV-positive cervical carcinoma cell lines CaSki and C33A served as positive and negative controls, respectively. Purified DNA concentration was quantified using the Qubit dsDNA HS Assay kit (Thermo Fisher Scientific).

### Trio-primer sets for pre-sequencing PCR viral enrichment

We designed primers for viral enrichment based on the HPV16 whole genome reference sequence (accession number K02718) from the Papillomavirus Episteme (PaVE) database^43^ **(Table 2)**. The Integrated DNA Technology (IDT) and the NCBI Primer-BLAST software were used to design three primer sets (**Fig. 1a and b**): Primer set 1- Near Full-length PCR primer set abbreviated **L_1_**, which generated long amplicons that span up to 7.7kb of the HPV16 genome. This primer set amplifies any intact or long HPV16 DNA sequences in a sample. Primer set 2- tiling-path primer sets abbreviated **T_1_**, **T_2_** (adopted from Yusuke Hirose et al., 2019)^44^, and **T_3_** give overlapping amplicons of approximately 2.1 kb, 3.9 kb, and 2.6 kb in length, respectively. This set of primers helps to enrich fragmented HPV16 DNA in a sample. Primer set 3- the junction primer set, abbreviated U1, amplifies the split region of HPV16 when it is linearized and integrated into the host. To assess the sensitivity of our primers, we performed serial dilutions of DNA extracted from the CaSki cell line, which is known to contain integrated HPV16.

### Polymerase chain reaction (PCR) for HPV16 components and conditions

DNA samples were amplified using specific primers and a high-fidelity PCR kit (New England Biolabs) on an Eppendorf Vapo Protect MasterCycler Pro system. Each 50 μl PCR reaction typically included 2 μL sample template DNA, 10 μM forward primers (2.5 µl), 10 μM reverse primers (2.5 µl), 10 mM dNTPs (1 µl), 5X Q5 Reaction Buffer (10 µl), (Nuclease-Free Water (31.5 µl) and Q5 High-Fidelity DNA Polymerase (0.5 µl). PCR amplification was performed for 40 cycles in the thermocyclers under the following conditions. All reactions were performed with human genomic C33A negative control DNA and Caski HPV16 positive control. The cycling conditions for each primer set were optimized as follows: The **U1** primer set underwent an initial denaturation at 98 °C for 90 s, followed by 40 cycles of denaturation at 98 °C for 10 s, annealing at 65 °C for 20 s, extension at 72 °C for 60 s, and a final extension at 72 °C for 2 minutes. Similarly, the **T1** primer set had an initial denaturation at 98 °C for 90 s, followed by 40 cycles of denaturation at 98 °C for 10 s, annealing at 66°C for 20 s, extension at 72 °C for 60 s, with a final extension at 72 °C for 2 minutes. For the **T2** primer set, the cycling conditions were as follows: an initial denaturation at 98 °C for 90 s, followed by 40 cycles of denaturation at 98 °C for 10 s, annealing at 65 °C for 20 s, extension at 72 °C for 5 minutes, with a final extension at 72 °C for 2 minutes. The **T3** primer set underwent an initial denaturation at 98 °C for 90 s, followed by 40 cycles of denaturation at 98 °C for 10 s, annealing at 64°C for 20 s, extension at 72 °C for 75 s, and a final extension at 72 °C for 2 minutes. Finally, the **L1** primer set underwent an initial denaturation at 98 °C for 90 s, followed by 40 cycles of denaturation at 98 °C for 10 s, annealing at 65 °C for 20 s, extension at 72 °C for 5 minutes, with a final extension at 72 °C for 2 minutes.

### Library preparation and Nanopore WGS of HPV16

The PCR amplicons were amplified separately for each primer pair. They were then pooled for each sample, cleaned using in-house PCR cleanup magnetic beads, and prepared for ONT sequencing using the Oxford Nanopore Rapid Barcoding kit (SQK-RBK110.96, Oxford Nanopore). Briefly, 9 μl of template DNA (ranging from 50 to 400 ng) was mixed with 1.4 μl of Rapid Barcodes (RB01-96, one for each sample), and the mixture was incubated in a thermocycler at 30°C for 2 minutes, followed by 80°C for 2 minutes, and then cooled to 4°C. The barcoded samples were pooled, and SPRI magnetic beads were added for bead clean-up in an equal volume. Beads were washed twice in 500 μl of 80% ethanol, and DNA was eluted in a 45 μl elution buffer. After pelleting the beads on a magnet, 45 μl of eluate containing the DNA library was collected, and the concentration of the library was measured using a Qubit fluorometer. Subsequently, 11 μL of the library was combined with 1.4 μL of Rapid Adapter F (RAP-F), mixed gently by flicking the tube, and then spun down. The mixture was incubated at room temperature for 5 minutes. The flow cell primer was prepared by mixing 30 μl Flush Tether (FLT) and 1170 μl Flush Buffer (FB) for loading onto the flow cell. For sequencing, 37.5 μl Sequencing Buffer (SQB), 25.5 μl Loading Beads (LB), and 12 μl of the DNA library were combined, and 75 μl of the library was loaded onto the Nanopore R9.4.1 flow cell for sequencing using a MinION sequencer.

### Base calling and demultiplexing of ONT sequences

Raw data (FAST5 files generated) from the Nanopore sequences were base-called and demultiplexed using Guppy 6.0.1 to FASTQ using the command for base calling with the R9.4.1l: guppy_basecaller -x "cuda:0" -c dna_r9.4.1_450bps_sup.cfg --input_path ./fast5 --save_path guppy6_sup_out/ --compress_fastq --enable_trim_barcodes --barcode_kits SQK-RBK110-96

### *De Novo* Assembly of HPV16 genomes

The HPV16 genome assembly was performed using Canu version 2.2^45^, which specializes in assembling long-read sequences. HPV16 fastq files for each sample were concatenated and used in genome assembly. For the de novo assembling process, we set Canu parameters as follows: ‘./canu readSamplingCoverage=100 -p sampleID -d sampleID minReadLength=500 minOverlapLength=250 genomeSize=8k -nanopore-raw sample.fastq.gz’. This process involved three phases: correction, trimming, and assembly, which improved base accuracy, removed low-quality regions, and generated contigs, consensus sequences, and alternate paths graphs. Finally, the assembled HPV16 genomes underwent manual correction and polishing using BioEdit v7.7.1.0 software.

### HPV16 reference-based variant calling pipeline

To analyze HPV16 sequences, we aligned long-read sequences to the HPV16 reference genome using Minimap2 with the command minimap2 -ax map-ont -a ./HPV16REF.fa Sample.fastq.gz > Sample.sam. The resulting SAM file was converted to a BAM file for efficient storage using samtools view -bS Sample.sam > Sample.bam. The BAM file was then sorted with samtools sort -O BAM Sample.bam -o Sample_sort.bam, and an index was created using samtools index Sample_sort.bam.

This pipeline effectively transforms raw sequencing data into a sorted and indexed BAM file, ready for variant calling and further analysis.

### Variant calling with PEPPER-Margin DeepVariant

To perform high-confidence variant calling from Oxford Nanopore Technologies (ONT) long-read data, we employed the PEPPER pipeline (version 0.8) using a Singularity container (pepper_deepvariant_r0.8.sif)^29^. Reads were aligned to the HPV16 reference genome (GenBank: K02718.1), and variant calling was conducted using the call_variant subcommand with the -- ont_r9_guppy5_sup model, optimized for super-accurate basecalled ONT data. The input BAM file (sorted BAM file *CaSki_HPV16h_sort.bam*) and reference genome HPV16REF.fa were processed using the specified pipeline parameters. Stringent filtering criteria were applied to improve precision and reduce false positives, including --pepper_snp_q_cutoff 10, --pepper_indel_q_cutoff 30, -- pepper_min_indel_baseq 20, --pepper_candidate_support_threshold 60, and -- pepper_indel_candidate_frequency_threshold 0.85. Additional thresholds were set for alignment quality and allele frequency: --pepper_insert_p_value 0.26, --pepper_delete_p_value 0.26, -- pepper_min_coverage_threshold 50, --pepper_min_mapq 44, --indel_min_af 0.15, and -- pepper_snp_p_value 0.000. Haplotype tagging was enabled via --use_pepper_hp, and phased BAM output was disabled using --skip_final_phased_bam. Variant calls were generated in compressed VCF format for downstream analysis.

A final filtering step was applied using bcftools to further enhance variant confidence, retaining only variants with a variant allele frequency (VAF) of 0.27 or higher. This threshold was chosen to minimize false positives while preserving true positive calls relevant to HPV16 genome analysis. The filtering was executed using the command bcftools view -i ’FMT/VAF[*]>=0.27’ -Oz -o Filtered_PMDV.vcf.gz Input_PMDV.vcf.gz, resulting in a refined VCF file used for downstream analyses.

### Clair3 variant calling

Variant calling was performed using Clair3 (v1.0)^30^, a tool optimized for long-read sequencing data. Aligned CaSki HPV16 reads (CaSki_HPV16h_sort.bam) were mapped to the HPV16 reference genome (GenBank: K02718.1). Clair3 was executed in a Singularity container (clair3_latest.sif), ensuring consistent deployment across computational platforms. The variant calling command included a model trained for Oxford Nanopore Technologies (ONT) data (*r941_prom_sup_g5014*), with 32 CPU threads allocated for parallel processing. The analysis was executed with parameters set to include all chromosomes (--include_all_ctgs), allow for long indels (--enable_long_indel), and a minimum SNP allele frequency of 7.5% (--snp_min_af 0.075). A minimum coverage threshold of 2 (-- min_coverage 2) was established, and variant calling was enabled at both the head and tail of sequences (--enable_variant_calling_at_sequence_head_and_tail). The chunk size was set to 8000 bases (--chunk_size 8000) for more efficient processing. All other parameters were maintained at their default settings, including those for haploid sensitivity, removal of intermediate directories, and use of the specified model path (--model_path).

With Clair3, variant calls were generated from both the full-alignment and pileup models. These two outputs were merged and filtered using a custom script to obtain a high-confidence set of variants. The merging process prioritized variants from the pileup VCF, including full-alignment variants only if they were not present in the pileup output. After merging, variants were filtered to retain only those that passed standard quality control (FILTER=PASS), had a genotype quality (GQ) score of at least 5, and did not contain duplicates. This strategy facilitated the integration of high-quality variant calls while minimizing noise, ensuring a reliable call set for HPV16 genome analysis.

### Establishment of an HPV16 truth set of variants

To establish proper control of HPV16 variants, also referred to as the ‘truth set’, we reprocessed publicly available short-read Illumina and in-house long-read Oxford ONT sequencing data derived from the CaSki cell line. The Illumina FASTQ files (European Nucleotide Archive accession: ERP111061) were initially aligned to both the human reference genome (GRCh38/hg38) and the HPV16 reference genome (GenBank: K02718.1) using HISAT2 (version 2.1.0) as described in the original study^46^. We independently realigned the same short-read data to the HPV16 reference genome using BWA-MEM2 (v2.2.1) for our analysis. Long-read ONT data generated in-house were processed using the same pipeline. Following alignment, we used SAMtools for sorting and indexing, GATK’s MarkDuplicates tool to mark PCR duplicates, and GATK’s AddOrReplaceReadGroups tool to assign read group information. Variant calling was performed using GATK HaplotypeCaller. The resulting VCF files from both platforms were merged, filtered to remove low-confidence sites, and manually curated against previously reported CaSki HPV16 Sanger variants^47^. This variant set served as a ground truth for benchmarking and concordance assessments of Clair3 and PEPPER.

## Statistical Analysis

The variant calls were analyzed using R (version 4.3.1; The R Project for Statistical Computing). Data were reported as means or medians for discrete and continuous variables, with counts expressed as percentages of the total. To ensure reliable sequencing across the low-coverage-prone region spanning positions 2,168 to 5,165, a minimum depth of 50× is typically recommended. In our dataset, the median read depth within this region was 7,026, indicating that our data exceeded the recommended coverage by approximately 140-fold and could be downsampled accordingly without compromising variant detection. Genomic benchmarking of variant callers is crucial for evaluating the concordance between ground-truth variants and called variants, as well as for identifying true positives (TPs), false negatives (FNs), and false positives (FPs). SNPs and indels were compared against our in-house “truth set” using a custom R script, classifying each variant as true positive (TP), false positive (FP), or false negative (FN). The script also computed key performance metrics, including precision (positive predictive value [PPV]), recall (sensitivity), and F1-score (harmonic mean of precision and recall), using the following formulas: Precision = True Positives / (True Positives + False Positives) (1) Recall = True Positives / (True Positives + False Negatives) (2) F1-Score = 2 x (Precision x Recall) / (Precision +Recall) (3)

### Phylogenetic analysis

Polished 12 clinical patient samples fasta sequences generated with Canu and HPV16 variant lineages and sub-lineages reference sequences were manually concatenated and subjected to Molecular Evolutionary Genetics Analysis version 11 (MEGA 11) software for pairwise and multiple sequence alignment. We analyzed the resulting Newick tree in the Chiplot Version 2.6.1 online tool.

## Ethical statement

Written informed consent was obtained from all participants. Ethical approvals were obtained from the Institutional Research and Ethics Committee (IREC) of Moi Teaching and Referral Hospital (MTRH) and Moi University, Eldoret, Kenya (reference number IREC/371/2022), as well as from the Institutional Review Board (IRB) of Brown University, Providence, Rhode Island, USA (IRB-ID: 2023003553).

## Data availability

All sequencing data has been submitted to SRA (#pending)

## Data Availability

All data produced in the present study are available upon reasonable request to the authors

## Acknowledgements

The authors are grateful to the nurses and study participants at the Cervical Cancer Screening Program at Moi Teaching and Referral Hospital in Eldoret, Kenya, for their hard work, dedication, and generosity in supporting this research. We thank our collaborators in Rwanda, including Gad Murenzi and his team, for providing the two clinical samples used in this study.

## Author’s contributions

Conceptualization and design of the study were led by M.K.T., D.G., C.I.O., A.C.E, R.A.K., and J.A.B. Optimization of the protocol M.K.T., D.G., C.I.O., R.M.C., A.A.F., S.T.H., K.N., and J.A.B. Data acquisition M.K.T., S.T.H., I.E.K., R.M.C., S.C., K.M., A.A.F., D.G., A.C.E, C.I.O., R.A.K., and J.A.B. Led the parent studies investigations P.K.T., P.M.I, P.J. L., A.M.M., R.T., D.R.B., O.E.O., S.C., A.C.E, R.A.K and J.A.B. Field work K.M., P.K.T., P.M.I, and O.E.O. Data analysis and interpretation M.K.T., D.G., C.I.O., R.M.C., A.A.F., S.T.H., K.N., R.A.K., and J.A.B. Writing original draft M.K.T., D.G., C.I.O., R.A.K., and J.A.B. Drafting and editing the article M.K.T., S.T.H., I.E.K., R.M.C., K.M., A.A.F., D.G., A.C.E, K.N., C.I.O., P.K.T., P.M.I, P.J. L., A.M.M., R.T., D.R.B., O.E.O., S.C., R.A.K and J.A.B. Funding Acquisition R.T., D.R.B., O.E.O., S.C., A.C.E, R.A.K, J.A.B., O.E.O., and P.J. L. Supervised R.A.K., and J.A.B.

All authors contributed to the writing and editing of the manuscript and approved the final version before submission.

## Corresponding author

Correspondence to Jeffrey A. Bailey.

---

**Supplementary Figure 1.**
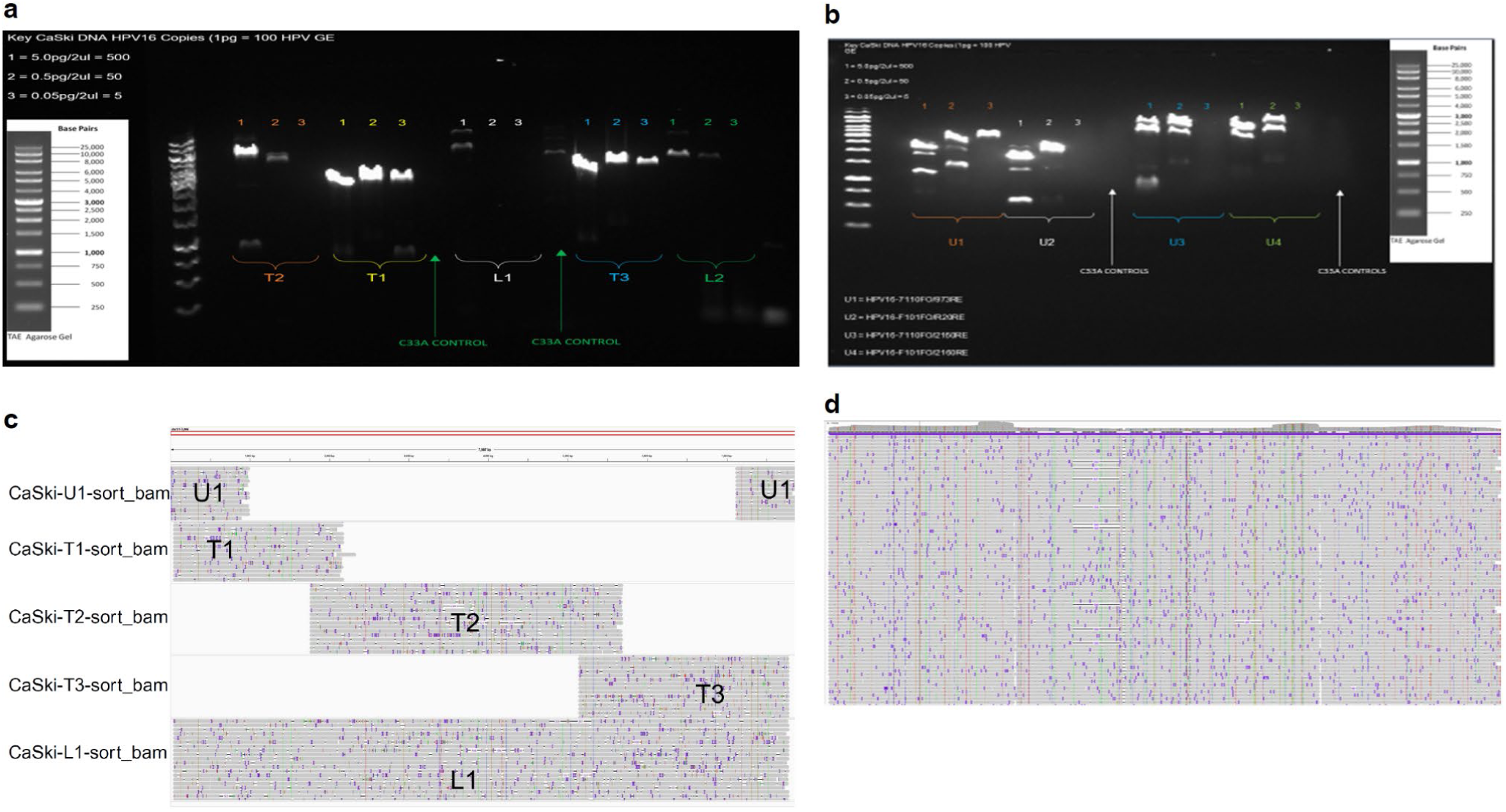
| Sensitivity of primer pairs for HPV16 whole-genome amplification. **(a)** Sensitivity testing of tiling primer sets T (yellow), T2 (orange), T3 (blue), and near full-length primer sets L1 (white) and L2 (green) using CaSki DNA at input of 5 PG (∼500 copies), 0.5 pg (∼50 copies), and 0.05 pg (∼5 copies) per 2 μL). **(b)** Evaluation of junction primer pairs spanning the HPV16 circular genome terminus: U1 (yellow), U2 (white), U3 (blue), and U4 (light green) across the same DNA input levels. **(c)** IGV view of bead-cleaned PCR products from U1, T1, T2, T3, and L1, showing expected amplicon coverage. **(d)** IGV visualization of pooled amplicons from all primer sets aligned to the CaSki HPV16 genome relative to the reference.

**Supplementary Figure 2.**
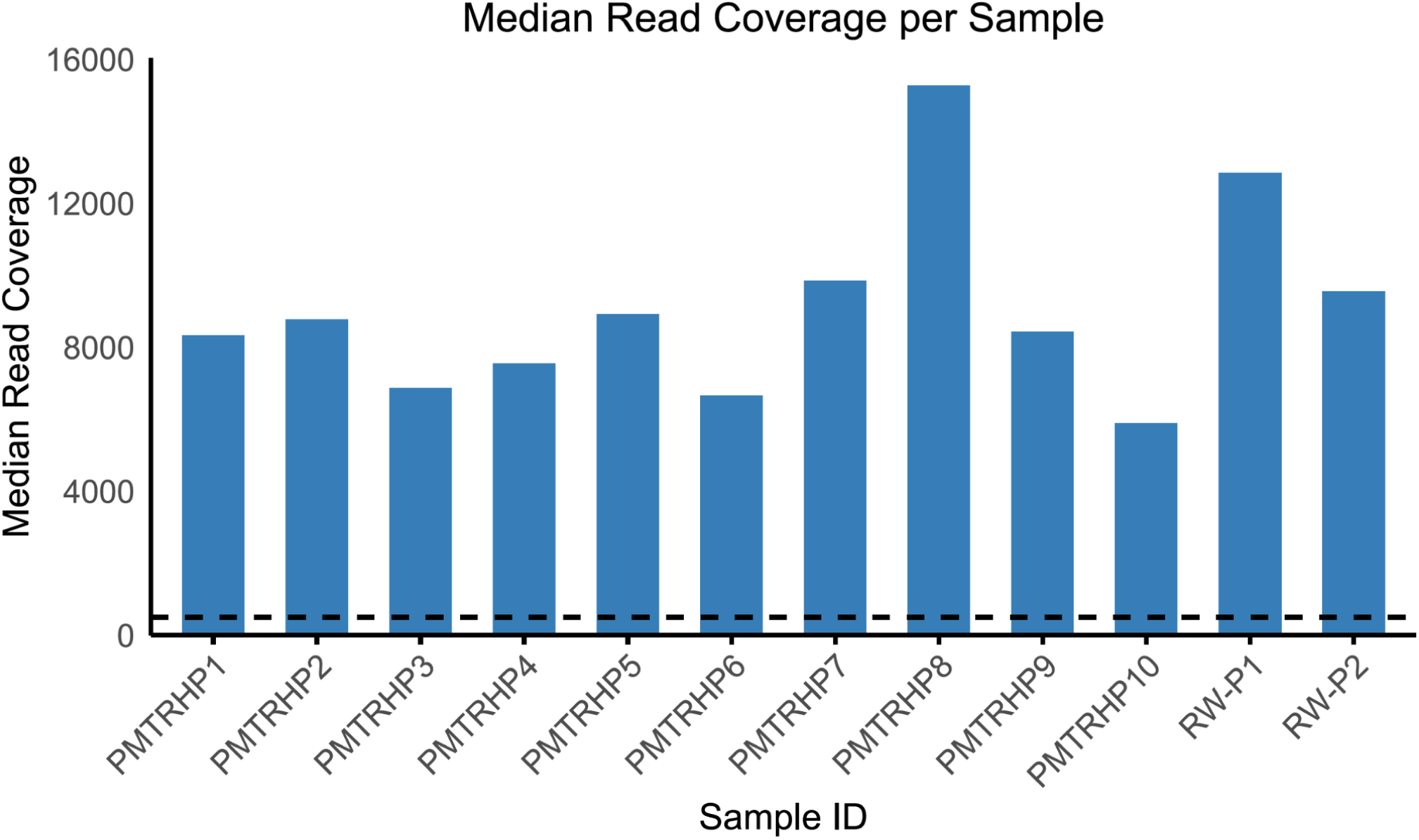
| Median read coverage per clinical sample. Bar plot depicting the median read coverage for each sample (n=12) obtained from long-read sequencing. A reference threshold of 500 median reads (dashed red line) shows that all samples obtained adequate coverage for further analyses.

**Supplementary Figure 3.**
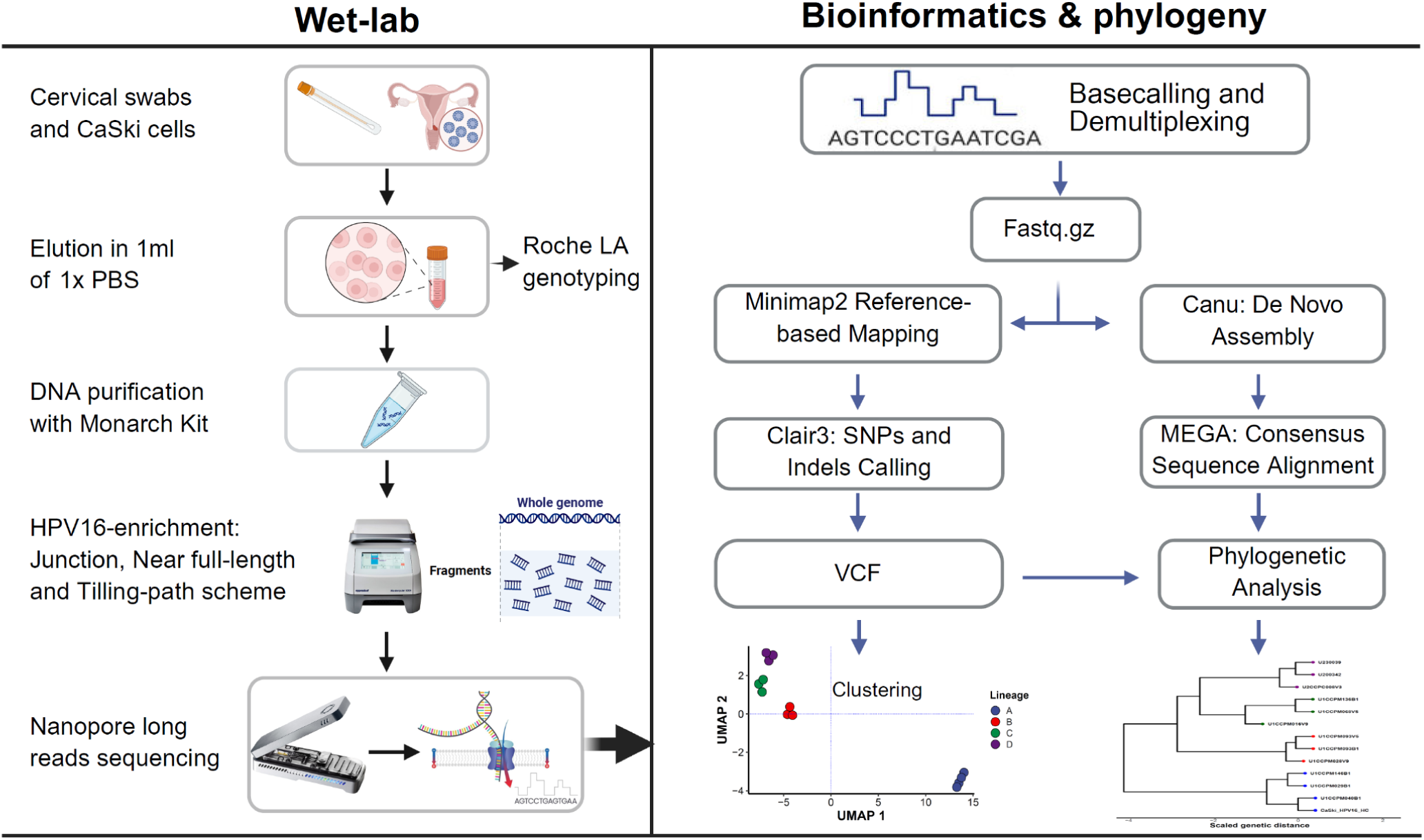
| HPV16 WGS and analysis pipeline. Left panel (wet-lab): DNA from the HPV16-positive CaSki cell line (used in method development) and cervical swabs from 12 women (validation cohort: aged years, NCI-funded Rwandan and Kenyan studies) were extracted, purified (Monarch kit, NEB), and sequenced on ONT to generate raw FASTQ reads. Clinical specimens were initially genotyped using the Roche Linear Array. Right panel (bioinformatics & phylogeny): ONT reads were base-called and demultiplexed, then aligned to HPV16 using Minimap2. The reads were sorted and indexed with SAMtools, and variants were called with Clair3, visualized in IGV. De novo assemblies (Canu) were aligned in MEGA for phylogenetic analysis.

